# Intracortical microstimulation evokes diverse electrode-specific phosphenes in the early visual cortex of blind individuals

**DOI:** 10.64898/2026.10.05.26364412

**Authors:** Leili Soo, Alfonso Rodil Doblado, Fabrizio Grani, Rocio Lopez Peco, Adrián Villamarin-Ortiz, Roberto Morollón Ruiz, Dorota Wacławczyk, Ione Fine, Pablo Gonzalez-Lopez, Cristina Soto-Sánchez, Arantxa Alfaro, Eduardo Fernandez

## Abstract

**Purpose:** Electrical stimulation of the occipital cortex evokes visual percepts known as phosphenes. To date, most human studies have examined phosphenes elicited by large surface or depth electrodes. These differ considerably from the small penetrating intracortical microelectrodes capable of stimulating highly localized cortical populations that are the basis of current next-generation devices. Here, we provide a detailed characterization of phosphenes evoked by such intracortical microelectrodes.

**Methods:** As part of the *CORTIVIS* clinical trial, a Utah Electrode Array with 100 penetrating electrodes was implanted in the early visual cortex of four blind volunteers. Participants completed drawing and description tasks in response to single-electrode stimulation. Two participants also performed forced-choice tasks to determine whether phosphenes elicited by different electrodes could be reliably discriminated.

**Results:** Single-electrode stimulation reliably evoked phosphenes in all participants. Most percepts appeared in the contralateral visual field, although two participants reported near-midline ipsilateral percepts. Phosphenes ranged from simple dots to elongated, textured, circular, or otherwise idiosyncratic forms. One participant reported distinguishable phosphene colors, which depended on electrode and amplitude structure and varied across sessions. The two participants tested on forced-choice discrimination could successfully identify electrode-specific phosphenes across multiple sessions.

**Conclusions:** Intracortical microstimulation of the human visual cortex produces phosphenes with diverse, electrode-specific appearances that are recognizable across repeated trials. These findings indicate that penetrating microelectrodes often evoke more structured percepts than simple points or blobs, providing foundational evidence that electrode-specific encoding strategies will be necessary in future visual prosthetic devices.

## Introduction

It is estimated that around 43 million people live with severe visual impairment worldwide ^1^ and this impairment has profound physical, psychological, and social impacts, significantly affecting quality of life ^2, 3^. For many diseases, such as individuals with blindness due to optic nerve damage caused by glaucoma or age-related macular degeneration, current therapies remain largely ineffective ^4, 5^. One potential solution involves visual prostheses, which aim to restore functional vision by electrically stimulating surviving neural pathways to elicit visual percepts, known as phosphenes ^6^. Phosphenes can be induced through stimulation at various stages of the visual pathway, including the retina ^7–14^, optic nerve ^15, 16^, lateral geniculate nucleus (LGN) ^17–19^ and occipital cortex ^6, 20, 21^. Among these, cortical visual prostheses have the potential to benefit the largest number of patients, since they can restore vision in patients with damage to the retina and/or optic nerve ^22–25^.

Although significant progress has been made in the development of cortical prostheses ^21^, one key area of uncertainty is whether these devices can produce ‘usable’ phosphenes. Qualitative data about phosphene appearance is difficult, if not impossible, to obtain from non-human animal models so most data on this topic come from early clinical studies in humans^6, 26–28^.

Although early reports of electrical stimulation in the visual cortex producing light perceptions date back to the early 20^th^ century ^29, 30^, the first attempt to use these perceptions as prosthetic vision for the blind started in the 1950s ^20^. These early studies, which included both surface and larger depth electrodes, described phosphenes that varied in shape, size, color, and brightness. Variability in phosphene appearance seemed to depend on stimulation site, stimulation parameters, and individual participant factors, but variability across phosphenes was observed even for prosthetics using very similar technologies (Table 1).

**Table 1.**
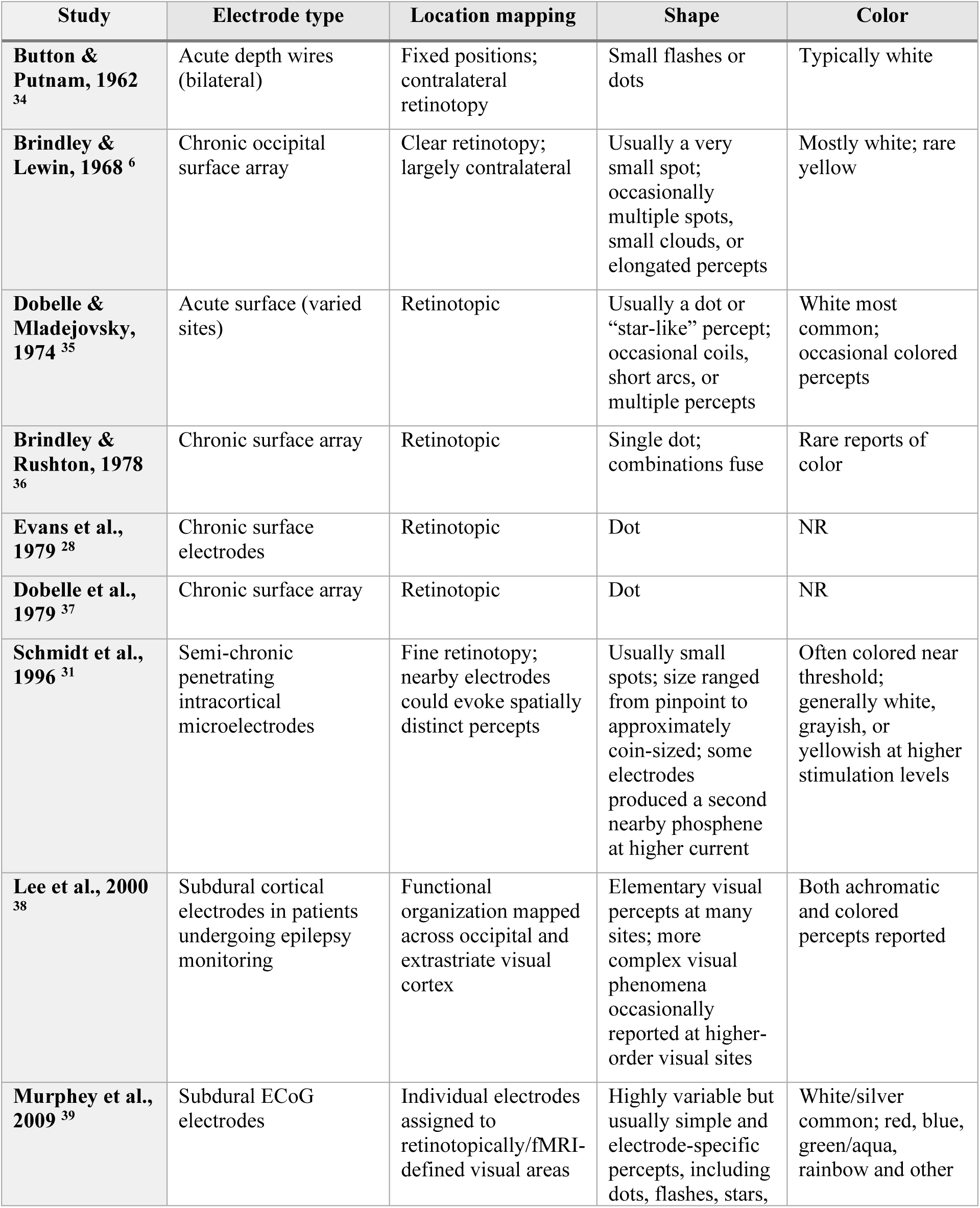
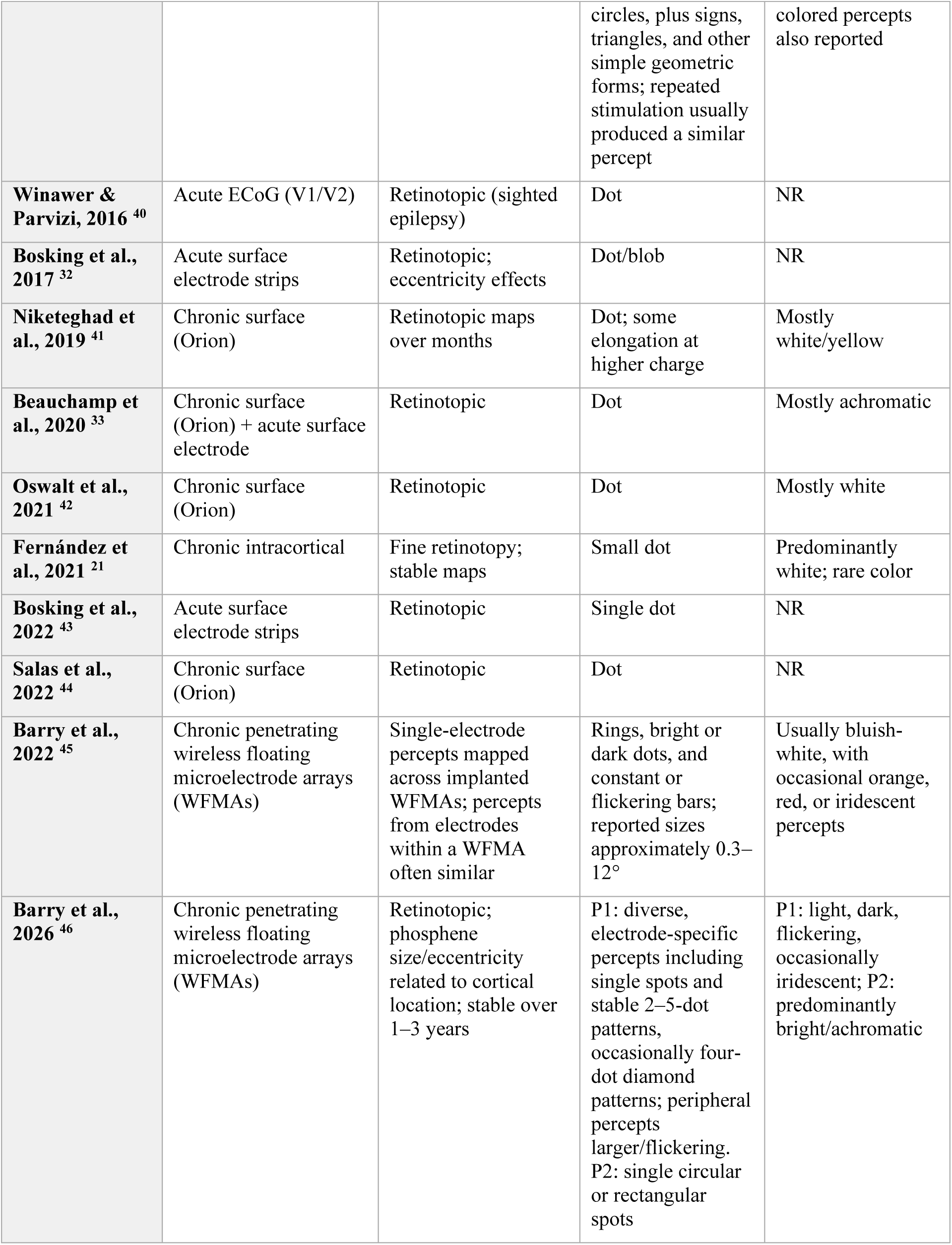
Human cortical stimulation studies characterizing percepts elicited by individual electrodes or stimulation sites.

Most existing literature does not systematically characterize the specific perceptual properties evoked by individual electrodes. The majority of studies report that single-electrode stimulation elicits focal light percepts, although it can less frequently induce complex geometries, such as elongated shapes or multiple phosphenes ^6, 31^. Additionally, the position where a phosphene is perceived typically aligns with the contralateral visual field relative to the stimulated hemisphere, reflecting the retinotopic mapping of visual cortex ^6, 21, 31^.

The size of phosphenes induced by electrical stimulation of single cortical electrodes initially increases with current amplitude and then saturates; additionally, percepts are larger for electrode sites farther from the foveal representation ^32^. In terms of appearance, percepts are most commonly described as achromatic or weakly colored (often white or yellow), though distinct color sensations have been reported, particularly near perceptual threshold ^6, 31^. Additionally, stimulation through multiple electrodes has enabled prosthetic implant recipients to perceive predictable patterns both via simultaneous stimulation ^6, 21^, and when using ‘rastering’ stimulation protocols which rely on stimulating electrodes in rapid temporal sequence ^33^. Some participants have described that with continuous stimulation the phosphenes disappear quickly, while others have report prolonged perception of phosphenes that persist even after stimulation has ended ^6, 31^.

While modern visual neuroprostheses increasingly rely on penetrating intracortical microelectrodes, detailed characterizations of the percepts elicited by single-electrode stimulation remain limited. In this study, we evaluate phosphenes elicited by individual electrodes of a high-density intracortical microelectrode array (Utah Electrode Array, UEA), focusing systematically on their fundamental features: location, shape, size, and color.

## Methods

### Participants

The study was approved by the Clinical Research Committee of the General University Hospital of Elche and registered in ClinicalTrials.gov (NCT02983370). All relevant ethical guidelines, including those for clinical trial regulations (EU No. 536/2014, repealed Directive 2001/20/EC), the Declaration of Helsinki, and European Commission Directives (2005/28/EC and 2003/94/EC), were adhered to. All participants provided written informed consent before any procedures were performed and were regularly reminded of their rights throughout the experiment. In accordance with ethical approval requirements, experimental periods were conducted consecutively so that each participant’s implant was removed before the next participant was implanted, ensuring no overlap in experimental periods.

Four blind human volunteers participated in this clinical study (three men). All participants had no light perception prior to the experiment, were in good general health, and had retinal or optic nerve damage, with no known damage within cortex. Further details are provided in Table 2. Throughout the six-month experimental period, each participant underwent various microstimulation protocols five times per week, with each session lasting approximately four hours, including breaks as needed.

**Table 2.** Participant Demographics.

| Participant | P1 | P2 | P3 | P4 |
| --- | --- | --- | --- | --- |
| Age group at implantation | 56-60 | 61-65 | 61-65 | 26-30 |
| Diagnosis | Bilateral Optic Neuritis | Bilateral Optic Neuritis | Bilateral Optic Neuritis | Ocular trauma |
| Level of blindness | No light perception | No light perception | No light perception | No light perception |
| Duration of blindness | >10 years | 2–5 years | >10 years | <2 years |
| Spontaneous visual percepts | Occasional | None | None | Present |

\*Age and duration of blindness are presented in categories to reduce the risk of participant identification.

### Implant

Intracortical electrical stimulation was delivered via Utah electrode arrays (UEA), originally developed by Dr Richard A. Normann ^47^ and manufactured for human use as NeuroPort Electrodes by Blackrock Neurotech (Salt Lake City, UT, USA). The UEA is an intracortical microelectrode array designed for neural recording and stimulation that consists of a grid of tiny silicon needles, each containing a conductive electrode tip capable of recording electrical signals from or delivering stimulation to neurons. Electrodes are arranged in a 10x10 configuration (the outer four electrodes are not active, see Figure 1), with each electrode 1.5 mm in length, enabling penetration into the cortical surface. This array is commonly used in neuroprosthetic applications, brain-machine interface research, and cortical visual prosthesis studies, allowing high-resolution, localized access to neuronal activity in specific brain regions, such as the motor cortex or visual cortex ^48^.

**Figure 1.**
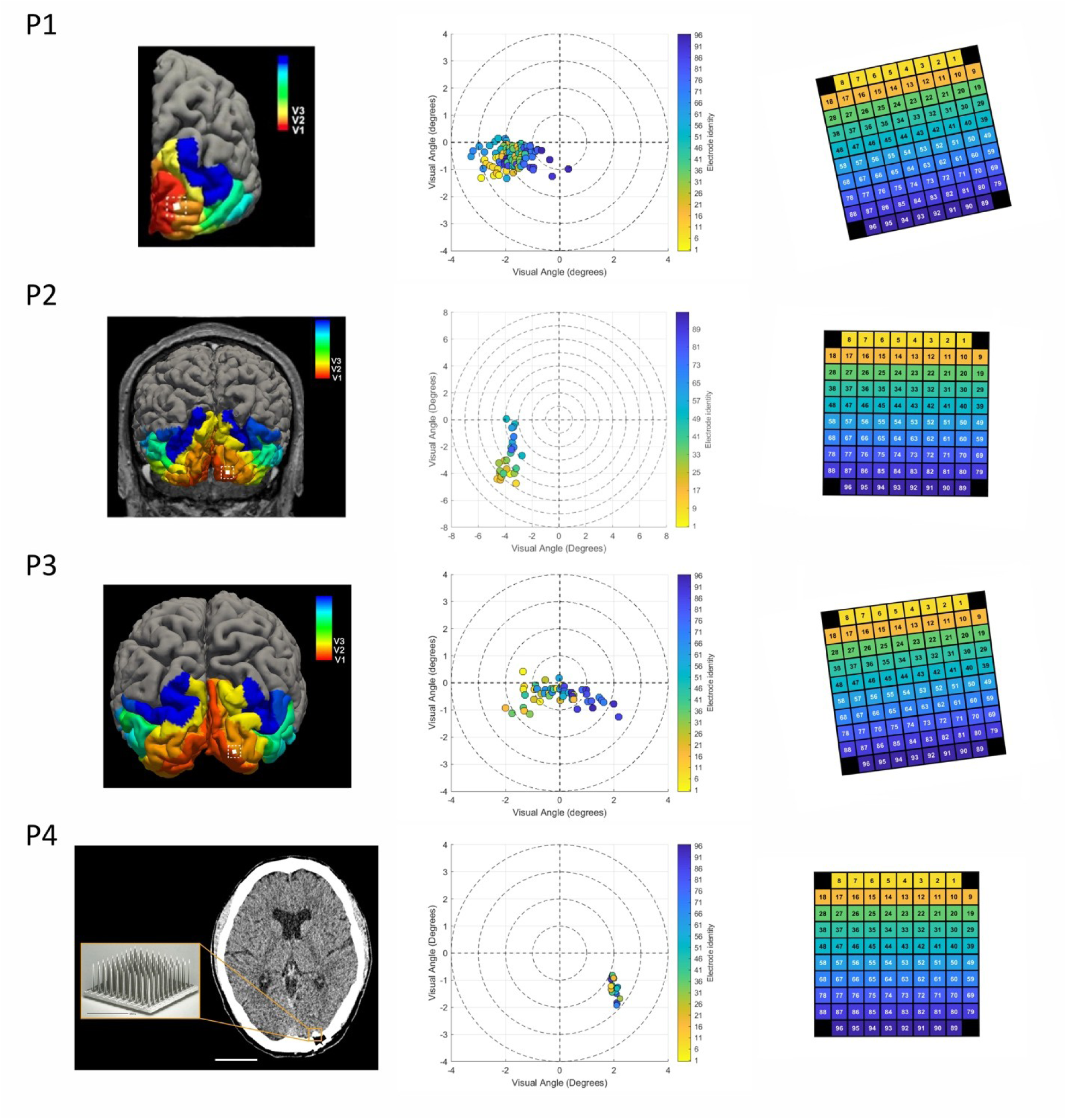
Implant locations and perceptual maps. **Left:** Implantation site is overlaid on predicted retinotopic map organization superimposed on the 3D reconstruction of each volunteer’s cerebral cortex. It was not possible to produce this mapping for the fourth volunteer. **Middle:** Locations of the perceived phosphenes, numbers indicate locations of electrodes in the Utah Array. Each dots indicates the mean reported phosphene location elicited by single-electrode stimulation, color-coded by electrode number. Note the change of scale across participants. **Right:** The Utah Array with color coded electrodes.

### Surgery and implant site

Before participation, volunteers underwent a comprehensive physical and mental evaluation as part of the pre-screening process. Upon completing all assessments, they were implanted with a UEA in the early visual cortex, for up to six months. The implant surgery for the first two participants was performed by a neurosurgeon following a standard procedure ^21^, while the surgeries for the last two participants were conducted using a robot-assisted implantation ^49^. Implantation sites were guided by transcranial magnetic or electrical stimulation (TMS or TES) protocol, where phosphenes were stimulated transcranially to identify the most effective locations for phosphene induction ^50^. The exact implantation sites were ultimately determined during the surgery by the neurosurgeon. We estimated the implant location via a predicted retinotopic map organization based on the Benson template ^51^ superimposed on a 3D reconstruction of the volunteer’s cerebral cortex that was obtained prior to surgery using MRI (Figure 1). The implantation sites were predominantly located in primary visual cortex (V1) or along the V1/V2 border. For the fourth participant, anatomical MRI could not be obtained due to clinical reasons; nevertheless, the array location was estimated to lie within V1. The first stimulation session was conducted at the hospital on the day after surgery.

### Stimulation parameters

Electrical microstimulation pulse trains were administered using a multichannel current stimulation system, Cerestim (Blackrock Microsystems Inc., Salt Lake City, USA) in the first participant and Ripple Neuromed Summit Explorer processor (Ripple Neuromed, Salt Lake City, USA) in the three following participants using custom-built proprietary stimulation software. Unless otherwise stated, electrical stimulation consisted of trains of 50 charge-balanced, cathodic-first biphasic pulses delivered at 300 Hz for 166 ms. Each pulse had a phase duration of 170 µs, an interphase interval of 60 µs, and a total pulse duration of 0.4 ms. Current amplitude was adjusted for each electrode to produce a bright, clearly visible phosphene within safe charge density limits.

### Phosphene Size, Shape, Brightness and Location

The size and position of phosphenes was reported through drawing the phosphene on a tablet which was mounted vertically on a board at a comfortable distance (∼50 cm). Before each stimulation, subjects were asked to maintain fixation on a tactile indicator glued to the middle of the tablet, and touch that fixation location with their right index finger. They were instructed that this represents the middle of their visual field. Participants were instructed to reproduce each percept as if it were viewed at arm’s length. The spatial extent of each drawing was converted to degrees of visual angle using the tablet dimensions and viewing distance.

To report relative brightness within the percept, participants shaded brighter regions with a higher density of pen strokes. The drawings were then processed offline: first, the sketch was converted into a grayscale image and smoothed using a Gaussian filter to interpolate stroke density and reconstruct the perceived brightness. Next, a threshold was applied to remove sparsely shaded artifacts generated when the stylus briefly contacted the tablet unintentionally; this step also sharpened edges that had been softened during Gaussian smoothing. Finally, the cleaned image was rendered using a white color map to enhance percept contrast for display and analysis.

### Phosphene Color

All participants were verbally asked to describe the color of the phosphenes elicited by intracortical stimulation. The first three participants described their phosphenes as “bright”, “white”, or “yellowish white”. As these participants did not report clearly distinguishable color categories, no further systematic color assessment was conducted in these participants.

P4 reported perceiving phosphenes in several distinct colors. To examine whether color percepts were stable when the same single electrode was repeatedly stimulated color reports were recorded during repeated single-electrode stimulation. Thirty-one electrodes were initially tested at two current amplitudes, 40 and 60 µA. Eight electrodes were excluded because color phosphenes could not be reliably elicited across repeated testing, leaving 23 electrodes for analysis. Each retained electrode was tested at both amplitudes across repeated testing blocks. On each trial, the participant verbally reported the perceived color of the phosphene. Reports were coded into the following categories: green, yellow, orange, white/silver, no phosphene, and not sure. The category “no phosphene” was used when no percept was reported, and “not sure” was used when a percept was detected but its color could not be reliably identified.

Color stability was assessed separately for each electrode and amplitude level. For this analysis, trials coded as “no phosphene” or “not sure” were excluded and electrode-amplitude pairs for which the participant reported a color on two or more trials were retained for analysis. For each retained pair, we identified the modal color and calculated modal consistency, defined as the proportion of valid repetitions in which the modal color was reported. Values ranged from 0 to 1, with higher values indicating greater color consistency across repeated testing. Because the number of repetitions for each electrode-amplitude pairing varied after excluding trials in which the response was “no phosphene”/“not sure”, modal consistency was interpreted descriptively rather than as a formal reliability estimate.

To test whether definite color reports were systematically associated with electrode identity and amplitude, permutation-based chi-square tests were conducted separately for the 40 and 60 µA conditions. For each amplitude, the observed Color × Electrode contingency table was compared with a null distribution generated by randomly permuting color labels across observations. This analysis tested whether definite color reports were randomly distributed across electrodes/amplitude levels or whether specific electrodes tended to elicit particular color reports. Cramér’s V was calculated as a measure of association strength.

### Electrode-specific variations in phosphene perception

Single-electrode phosphene-appearance stability was assessed in two participants, P3 and P4, using a n-alternative forced-choice (AFC) appearance-matching paradigm designed to determine whether repeated stimulation of a given electrode produced a stable and recognizable phosphene. P1 and P2 did not report sufficiently distinct single-electrode phosphene shapes to support appearance-matching tasks. During preliminary testing, electrodes that elicited distinct and reproducible percepts were selected. The participant was then trained until they reliably associated stimulation on a given electrode to a specific response key. Then, in each trial, an electrode was stimulated, and the participant selected the response key that best matched the phosphene elicited on that trial. Within each testing block, electrodes stimulation occurred in pseudorandom order. After each response, an auditory cue indicated whether the answer was correct or incorrect. The next stimulation was delivered after a 2-second delay, and no response time limit was imposed. Response times were not analyzed.

P3 completed two- and three-alternative forced-choice tasks. P4 completed two-, three-, and four-alternative forced-choice tasks. In the two-AFC task, participants selected between two predefined electrodes; in the three- and four-AFC tasks, they selected from three or four predefined electrodes, respectively.

## Results

### Phosphene Size, Shape and Location

The Utah Electrode Array (UEA) was implanted in the early visual cortex of the right hemisphere in all participants (Figure 1 left panel). For P1 and P2 phosphenes were always perceived in the contralateral visual field and the location of the phosphenes followed a retinotopic organization (Figure 1 middle panel). For P2 phosphenes elicited by single electrodes were large and had more complex shapes, making it challenging for the participant to consistently identify their central location. For P3 the phosphenes appeared in both visual fields. For P4 phosphenes were seen in the ipsilateral visual field. These phosphenes were close to fixation and are therefore interpreted as near-midline variability rather than a violation of retinotopic organization. For P4, spatially distinguishing between phosphenes from different electrode stimulations was difficult because they clustered closely together (within ∼1 degree).

Perceived phosphene shapes produced by single electrode stimulation varied substantially across participants (Figure 2). In P1 (Figure 2, P1), phosphenes most often appeared as pinpoint lights at arm’s length. In P2 (Figure 2, P2), electrodes that elicited phosphenes produced square-like percepts containing either vertical or horizontal internal lines. In P3 (Figure 2, P3), most electrodes evoked recognizable and distinct forms that the participant could reliably discriminate and reproduce. P4 reported that the visual field appeared red, and stimulation most commonly generated circular or semi-circular percepts (Figure 2, P4).

**Figure 2.**
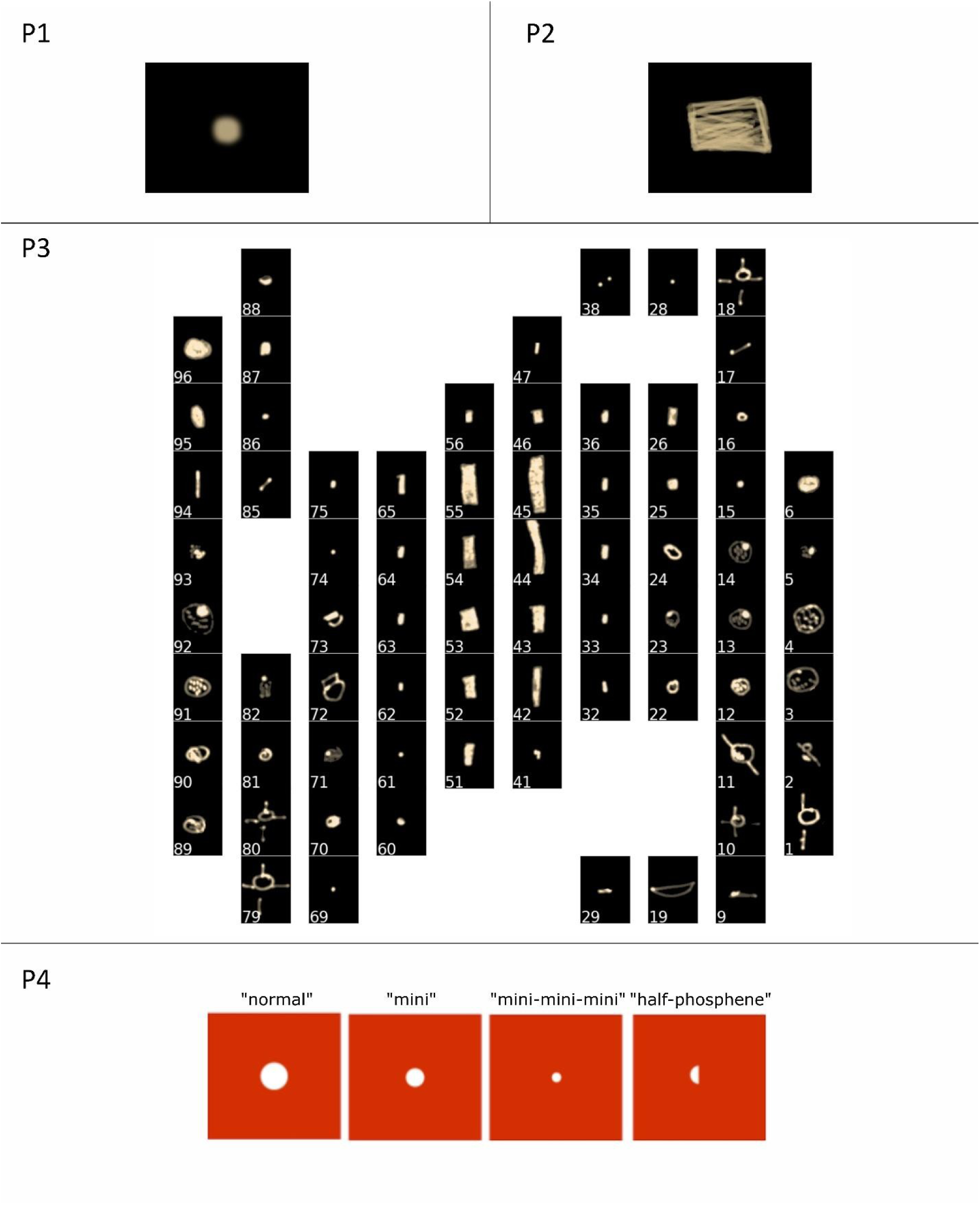
Representative phosphenes reported during single-electrode stimulation reconstructed from participant tablet drawings. Numbers indicate stimulated electrode IDs. P1 typically reported point-like percepts; P2 reported rectangular percepts with internal line structure; P3 reported a range of electrode-specific shapes; and P4 reported circular or partial circular percepts against a red visual background.

Phosphene sizes differed markedly between participants (Figure 3). P1 is not shown in the violin plots because the reported percepts were consistently point-like, with an estimated diameter of approximately 0.1° VA - substantially smaller than those of the other participants. P2 perceived predominantly rectangular phosphenes, with a consistently larger horizontal than vertical extent (0.91 ± 0.05° VA in width; 0.53 ± 0.06° VA in height). P3 who reported a diverse set of shapes, also reported a comparable diversity in size, with mean extents of 0.32 ± 0.25° VA in width and 0.32 ± 0.23° VA in height. P4 perceived phosphenes whose size clustered into discrete categories.

**Figure 3.**
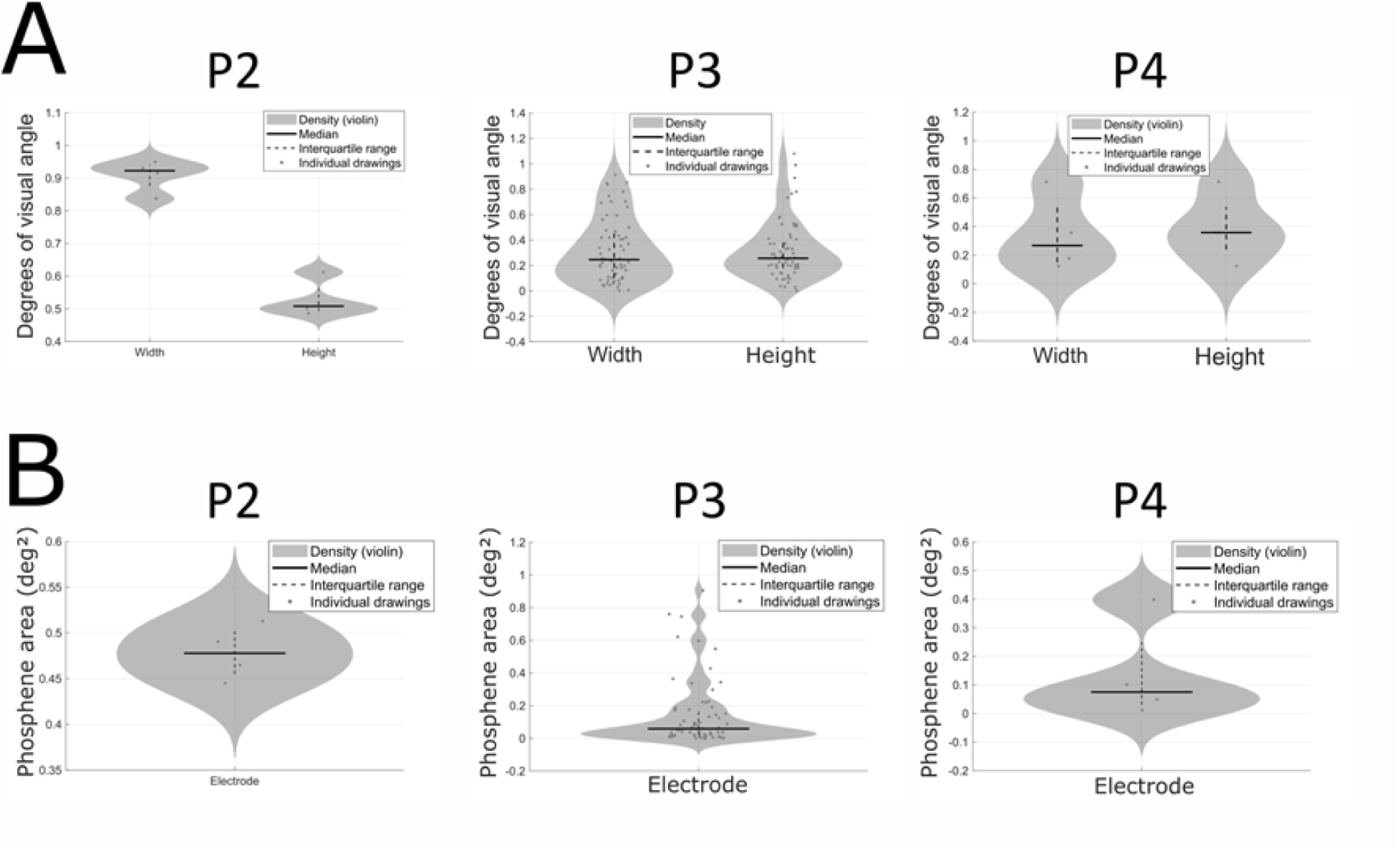
Phosphene sizes from single-electrode stimulation across participants. Note the difference in scale across participants. A) Phosphene extent in degrees of visual angle, shown separately for width and height. B) Phosphene area derived from the drawn phosphene shape. In all panels, violins represent kernel density estimates of the distributions, solid horizontal lines indicate medians, thick dashed lines denote interquartile ranges, and dots correspond to individual drawings.

Approximately circular phosphenes were described as “normal” (∼0.7° VA diameter), “mini” (∼0.4° VA diameter), or “mini-mini-mini” (∼0.12° VA diameter). P4 also reported “half” phosphenes, which were approximately semi-circular or elongated partial percepts measuring ∼0.4 × 0.12° VA.

### Phosphene Color

Three of the four participants reported white or yellowish white phosphenes. P4 reported phosphenes in several distinguishable colors, including white/silver, yellow, green, and orange. Some phosphenes were perceived as too small or brief for the participant to identify their color reliably. P4 also reported a consistent subjective relationship between color and brightness: white or silver phosphenes were described as the brightest, yellow and green percepts as intermediate, and orange phosphenes as the darkest. However, the relationship between stimulation current and perceived color was not straightforward; higher current did not consistently produce a shift toward brighter or more achromatic reports.

To assess color stability across repeated stimulation, color reports were analyzed for each electrode-amplitude pairing across repeated testing blocks. After excluding “no phosphene” and “not sure” responses from the modal-consistency analysis, 44 electrode-amplitude pairings had at least two repeated measurements. At 40 µA, mean modal consistency was 0.71 and median modal consistency was 0.75, meaning that the same color (out of four options) was reported on approximately 3 out of 4 repeated trials. At 60 µA, consistency was lower, with a mean modal consistency of 0.59 and a median of 0.50, meaning that the same color was chosen on approximately half the repeated trials. However, some electrodes were very consistently associated with a particular color: 12 of 44 electrode-amplitude pairs reached a modal consistency of at least 0.83, the same color was reported in at least five of six repeated trials.

Color reports were not randomly distributed across electrodes. Permutation-based chi-square tests showed a significant association between color category and electrode identity at both 40 µA, χ² = 109.17, p = 0.0004, Cramér’s V = 0.45, and 60 µA, χ² = 107.89, p = 0.0006, Cramér’s V = 0.44. Thus, although color percepts were only partially stable across repeated testing, there was a systematic relationship between electrode and phosphene color.

**Figure 4.**
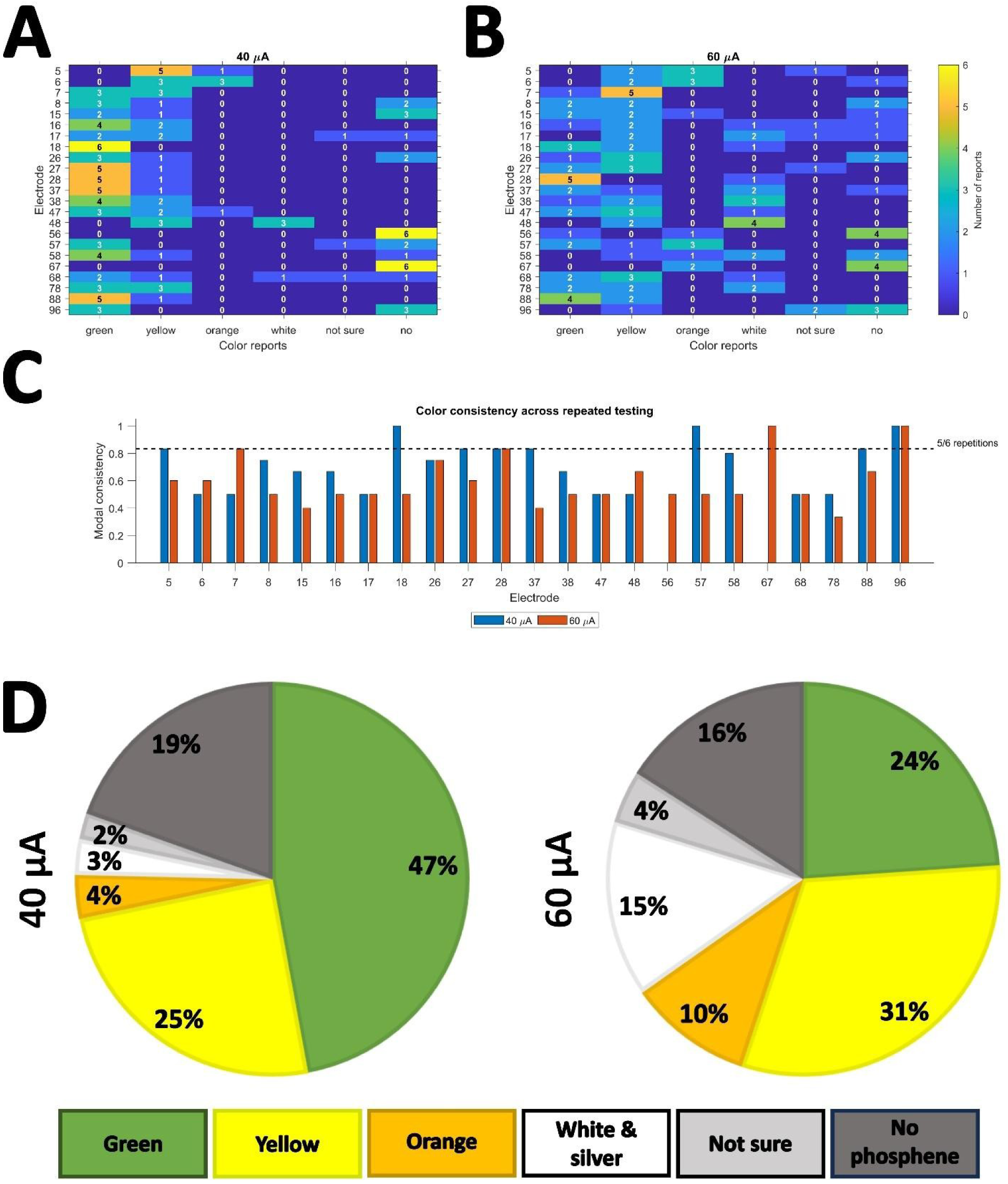
Color percepts elicited by single electrode intracortical stimulation for P4. **A–B**) Heatmaps showing the distribution of verbally reported phosphene colors across electrodes at 40 µA (A) and 60 µA (B). Each row represents one electrode, and each column represents one response category. Cell values indicate the number of reports for each color category across repeated testing sessions. The color scale indicates the number of reports. “No” indicates trials in which no percept was reported. **C**) Modal color consistency for each electrode-amplitude pair across repeated testing. For each electrode and amplitude, the modal color was identified after excluding “no phosphene” and “not sure” responses, and consistency was calculated as the proportion of valid repetitions in which that modal color was reported. The dashed line indicates a consistency of 5/6 repetitions. **D**) Overall distribution of color reports at 40 µA and 60 µA. Percentages indicate the proportion of reports assigned to each color category. “No phosphene” indicates trials in which no percept was reported, and “not sure” indicates trials in which a percept was detected but its color could not be reliably identified.

### Electrode-specific differences in visual percepts

Different electrodes elicited distinct and recognizable phosphenes - behavioral matching accuracy was well above chance for each n-AFC task (Figure 5). Across testing days, participants verbally reported that the same electrodes produced similar phosphene appearances, indicating qualitative across-day stability. For P3, the combined two- and three-AFC analysis included 240 stimulation trials across eight testing dates. Accuracy was 91.7% in the two-AFC task and 97.5% in the three-AFC task. Ten unique electrodes were tested, of which eight were assessed on more than one testing date. Electrode-level behavioral matching accuracy ranged from 85.0% to 100% (Figure 5A).

**Figure 5.**
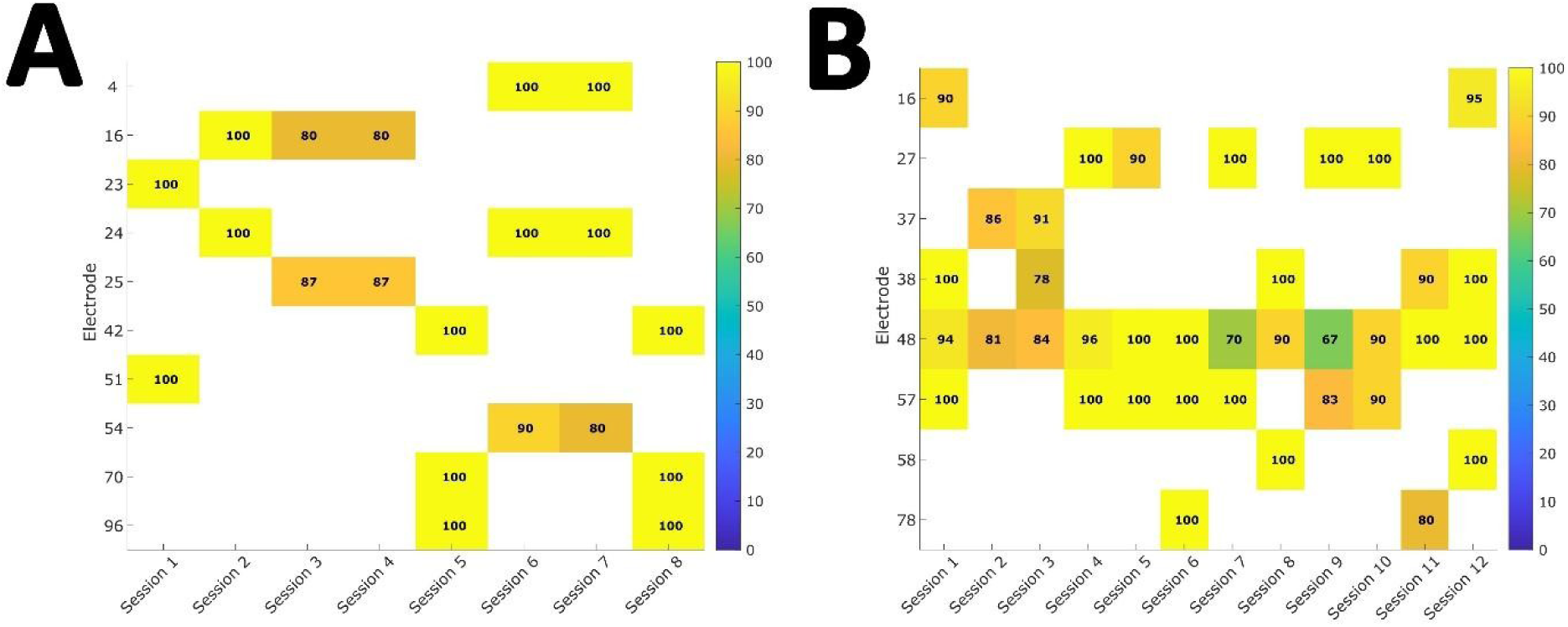
Cross-session consistency of phosphenes for P3 and P4. Behavioral matching accuracy is shown for each stimulated electrode across testing sessions in P3 (A) and P4 (B). Each cell represents the percentage of trials in which the participant correctly identified the electrode that had been stimulated. Values within colored cells indicate accuracy. White cells indicate that the electrode was not tested in that session.

For P4, the combined two-, three-, and four-AFC analysis included 940 stimulation trials from eight unique electrodes. Accuracy was 90.0% in the two-AFC task, 93.1% in the three-AFC task, and 87.9% in the four-AFC task. Electrode-level behavioral matching accuracy ranged from 86.8% to 100% (Figure 5B), although some trials produced non-matching responses.

Together, these results indicate that single electrodes can evoke recognizable electrode-specific phosphenes whose appearance was qualitatively stable across days and behaviorally discriminable within AFC testing sessions.

## Discussion

Intracortical stimulation of early visual cortex elicited reliable phosphenes with diverse perceptual appearances across four blind volunteers. We observed (i) predominantly contralateral phosphene locations, though phosphenes were reported ipsilateral near the midline reports in two volunteers, (ii) a wide range of reported shapes, from point-like percepts to elongated, textured, or otherwise idiosyncratic forms, (iii) evidence that selected electrodes could evoke recognizable electrode-specific appearances across repeated testing sessions in two participants, and (iv) color reports in one participant, which were dependent on both the electrode and the current amplitude, but only partial consistency across repeated testing sessions.

Together, these findings suggest that intracortical microstimulation can elicit phosphenes more complex than the simple “spots” or “light blobs” often emphasized in the literature. They also show that the stimulating electrode can have an important influence on perceived phosphenes, supporting the view that electrode-specific perceptual characteristics should be considered when designing stimulation strategies for visual cortical prostheses.

As expected from striate/early extrastriate retinotopy ^6, 21^, single-site stimulation typically yielded contralateral phosphenes. In two volunteers, some reports fell just into the ipsilateral field but remained close to fixation (∼3°). This small deviation is consistent with known uncertainty around the vertical meridian and with reports that receptive-field edges can show limited inter-hemifield border intrusions in humans and non-human primates ^52–55^. Several non-exclusive explanations are plausible. First, fixation estimation in blindness may involve proprioceptive anchoring and introduce a small constant bias ^56–58^. Second, stimulation near the V1/V2 border or close to the vertical meridian may activate neural populations with receptive fields that lie near, or partially straddle, the inter-hemifield boundary ^53, 54, 59^. Given the tiny absolute deviation relative to the ∼190–200° horizontal visual field, these ipsilateral reports are best viewed as near-midline variability rather than a violation of retinotopic organization.

We also observed clear between-electrode idiosyncrasies and between-participant differences in phosphenes. P1 reported percepts ranging from pinpoints to elongated shapes. P2 reported rectangular shapes across all electrodes. P3 reported the greatest diversity of clearly recognizable phosphene shapes and P4 reported simple circle- or half-circles which varied in size.

The reason for these individual differences remains unclear. They cannot be explained by duration of blindness alone: P1 and P3 had long-standing blindness, whereas P2 and P4 had more recent vision loss. Likewise, array location offers no clear explanation, as all implants were situated in similar anatomical regions (V1 or near the V1/V2 boundary). These observations suggest that phosphene shape reflects an interaction between cortical implantation site, local cortical architecture, electrode–tissue interface properties, stimulation parameters, and individual visual history, including the cause and duration of blindness.

Classical intracortical studies have noted that colored phosphenes are most common when stimulation is near threshold, with percepts tending toward white/gray/yellow as intensity increases ^31^. In our cohort, only one of four participants reported colored phosphenes, despite identical hardware and similar levels of stimulation relative to threshold compared to the other participants. This participant continued to perceive colored phosphenes at suprathreshold currents, although phosphenes tended to become more white or yellow at higher currents. This same individual also reported a persistent red visual background. Together, these observations suggest that a blind person’s ongoing internal visual state (e.g., deafferentation-related spontaneous visual phenomena such as Charles Bonnet–type hallucinations or visual-snow–like backgrounds ^60, 61^) might bias phosphene chromaticity, paralleling state-dependent effects seen in TMS, where color adaptation can modulate the color of TMS-induced phosphenes ^62, 63^. This argues for explicitly assessing a participant’s baseline “visual background” before stimulation sessions and considering adaptation/history effects when interpreting color reports.

Our results strongly suggest that a deeper understanding of phosphene generation will be needed to develop stimulation strategies that will yield useful prosthetic vision. For decades, clinical testing of electrical stimulation, at retinal and cortical levels, has progressed in parallel with simulated prosthetic vision (SPV) studies in normally sighted observers and in silico models. Classic and recent SPV papers show that many object-level behaviors (faces/letters/objects) can be supported with appropriate pre-processing, while navigation and obstacle avoidance benefit from scene simplification and head-scanning strategies ^64–67^. Most SPV work, however, has assumed pixelated vision, meaning regular grids of uniform, Gaussian “dots” (“scoreboard” models), a simplification explicit in foundational reviews and widely used in contemporary VR tasks ^67, 68^. Our results suggest that assuming a simple electrode-to-pixel mapping is unlikely to be sufficient ^69^. Instead, stimulation strategies may need to treat each electrode-specific phosphene as a distinct visual element, extending to cortical prostheses the logic of model-based and dictionary-learning approaches proposed for retinal prosthesis design ^70^.

In summary, we show that intracortical microstimulation of early visual cortex elicits phosphenes with diverse, electrode-dependent appearances in blind participants. In the two participants who completed appearance-matching tasks, the phosphenes elicited by electrodes were recognizable across repeated testing sessions with a high degree of accuracy. These findings challenge simple electrode-to-pixel assumptions and suggest that future cortical visual prostheses may need to incorporate electrode-specific perceptual features into stimulation and image-encoding strategies.

## Data Availability

All data produced in the present study are available upon reasonable request to the authors.

## Acknowledgements

The authors express their sincere gratitude to all four participants and their families for their dedication and involvement in this clinical study. We also thank the clinical and administrative staff at IMED Hospital Elche for their invaluable assistance, ongoing clinical care, and patient monitoring throughout the study.

## Commercial Relationships Disclosure

None.

